# Risk Factors for Low Humoral Response to BNT-162b2 In Hemodialysis Patients

**DOI:** 10.1101/2021.09.21.21263568

**Authors:** Rui Duarte, Cátia Figueiredo, Ivan Luz, Francisco Ferrer, Hernâni Gonçalves, Flora Sofia, Karina Lopes, Ana Vila Lobos

## Abstract

**Introduction:** Maintenance Hemodialysis (HD) patients are at higher risk of both infection and mortality associated with the new coronavirus 2. Immunization through large-scale vaccination is the cornerstone of infection prevention in this population. This study aims to identify risk factors for low response to the BNT-162b2 (Pfizer BioNTech) vaccine in a HD cohort.

**Materials and Methods:** Observational prospective study of a HD group followed in a Portuguese Public Founded Hemodialysis Center who received BNT-162b2 vaccination. Specific anti-Spike IgG was evaluated as arbitrary units per milliliter (AU/mL) on two separate occasions: 3 weeks after the first dose and 3 weeks after the second. IgG titers, Non-Responders (NR), and Weak-Responders (WR) after each dose were evaluated against risk factors that included demographic, clinical and analytical variables.

**Results:** Humoral response evaluated by IgG anti-Spike levels showed a strong correlation with Charlson comorbidity index (CCI) and intact parathormone (iPTH) after each inoculation (1^st^ dose: ρ=−0.64/0.54; 2^nd^ dose: ρ=−0.66/0.63, respectively; p<0.01 throughout). After completing both doses: 1) NR were associated with female sex (p<0.01), lower albumin and iPTH (p=0.01); 2) WR showed higher CCI, older age, lower iPTH and lower albumin (p=<0.01, p=0.03, p<0.01, p=0.05, respectively) and, consistently, associated with CCI over 8, age over 75, iPTH under 150 ng/L, female sex, dialysis vintage under 24 months and central venous catheter (CVC) over arteriovenous fistula (p=0.01, p=0.03, p<0.01, p=0.01, p=0.01, p<0.01, respectively). A binary regression model using CCI, sex (male) and CVC was statistically significant in prediction of WR after the 2^nd^ dose with OR (95% CI): 1.81 (1.06-3.08); 0.05 (0.01-0.65); 13.55 (1.06-174.18), respectively (p=0.01).

**Conclusion:** Older age, higher CCI, lower iPTH and albumin, CVC as vascular access and recent hemodialysis initiation (less than 2 years) associate with lower response to vaccination in our study. A higher comorbidity burden is suggested as a more significant surrogate marker for low immunogenicity rather than age alone. Identifying HD patients as a population at high-risk for low response to vaccination is essential for proper policy-making, facilitating the implementation of adequate and individualized contingency protocols.

**What is already known about this subject:**

- Maintenance hemodialysis patients have lower humoral response to BNT-162b2 COVID-19 vaccine when compared to the general population.
- Maintenance dialysis patients are at high risk of exposure to coronavirus 2 in addition to a more severe disease course.

**What this study adds:**

- We suggest Charlson commorbidity index, older age, intact parathormone, central venous catheter as vascular access and lower dialysis vintage as possible surrogate markers of immunogenicity in HD patients.
- There is a low humoral response after a single dose of the vaccine (50%) that can be increased after the second (86%).

**What impact this may have on practice or policy:**

- Strict Protocols for follow-up measures in HD patients, including closer humoral titers assessment, risk stratification, adequate isolation, and surveillance of symptoms might be necessary in order to improve this population survival/life expectancy.
- Screening HD patients, seroconversion rates may be improved by giving extra inoculations for patients at risk for low response.

## Introduction

Global immunization against severe acute respiratory syndrome coronavirus 2 (Sars-CoV-2) is the standard-of-care in preventing Coronavirus disease 2019 (COVID-19). Different vaccines, with different action mechanisms have been developed, namely BNT-162b2 (Pfizer-BioNTech)^1^ and mRNA-1273 (Moderna)^2^ – mRNA-based vaccines; ChAdOx1 nCov-19 (AstraZeneca)^3^ and Ad26.COV2.S (Johnson&Johnson/Janssen)^4^ – recombinant adenovirus vectors encoding Sars-CoV-2 spike glycoprotein.

The need for mandatory regular contact with health care services in maintenance hemodialysis patients (HD), coupled with worse disease severity and increased mortality risk, establish HD patients as a high-risk population.^5-7^ In this setting, international recommendations considered vaccination of HD population as a priority and the Portuguese government included it early in the national immunization plan, starting on February 2021. BNT-162b2 was the available and inoculated vaccine in our Nephrology center. Efficacy studies for this vaccine did not include HD patients,^1^ delaying scientific data on the subject. Currently, multiple studies using real-world evidence, through humoral response assessment and comparison of infection rates, recognized the HD population as less responsive to vaccination, when compared to the general population.^8-10^

Furthermore, antiviral immunogenicity in HD patients has been a long-known issue, particularly concerning antiviral vaccination, as illustrated by hepatitis B, with only 50 to 60% of patients exhibiting humoral response in the first studies when compared to over 90% in healthy individuals.^11,12^ To minimize this problem, new protocols were established involving follow-up antibody measurements, adjuvant and new vaccines development and repeated inoculations, improving seroconversion in HD patients to 80%.^13^

The identification of patients at risk for low response to vaccination is vital in the implementation of proper contingency protocols, mostly revolving around proper isolation, testing and symptom vigilance. Moreover, and similarly to Hepatitis B, repeated inoculations might be appropriate in the HD subgroup whose vaccination does not elicit a response after the conventional vaccination schedule.

Therefore, this prospective study conducted in a Portuguese Public Founded Hemodialysis Center, aimed to describe humoral response to the BNT-162b2 vaccine in a cohort of HD patients and identify possible risk factors that might be used to identify those at high risk for inadequate response to vaccination.

## MATERIALS AND METHODS

The observational study prospectively enrolled forty-six HD patients who were scheduled to receive two doses of BNT-162b2 with a three-week interval, in accordance with pharmaceutical guidelines for administration.

The assessment of vaccination response was done as part of the internal policy of the center’s contingency protocol and informed consent of each individual patient was obtained regarding the access and use of sociodemographic, clinical and laboratorial information for scientific. Blood collection and analysis was made at two distinct phases: 1) Three weeks after the first dose was administered and 2) Three weeks after the second. Because the recommended dosing interval is three weeks, the first collection was made coincidentally to the second dose administration.

We performed the measurement of IgG anti-Spike for humoral response and IgM, both anti-Nucleocapsid and anti-Spike, in order to track down patients who could have contacted with the virus. Titers were measured as arbitrary units per milliliter (AU/mL) using chemiluminescence. Response was considered significant for titers superior to 1 AU/mL for both the 1^st^ and 2^nd^ doses.

Age, sex, presence of diabetes, Charlson comorbidity index (CCI), dialysis vintage, vascular access, albumin, intact parathormone (iPTH) and C-reactive protein (CRP) were established as the proposed risk factors. This data was collected from medical records and routine monthly laboratory analysis.

### Statistical analysis

Statistical analysis was carried out using Microsoft Excel 2016 and IBM SPSS Statistics 25 software.

Variables used in this study are summarized in table 1. Three humoral variables were defined: 1) IgG anti-spike levels; 2) Non-Responders (NR); 3) Weak-Responders (WR) corresponding to an IgG below percentile 25. Risk factors were also divided in continuous and categorical, when appliable.

**Table 1:** Continuous and categorical variables included in the study divided in humoral related and risk-factor related.

| Continuous | Categorical |
| --- | --- |
| **Humoral response associated variables** |  |
| IgG anti-Spike titers 1<sup>st</sup> dose (AU/mL) | 1<sup>st</sup> dose NR (<1AU/mL) |
| IgG anti-Spike titers 2<sup>nd</sup> dose (AU/mL) | 2<sup>nd</sup> dose NR (<1AU/mL) |
|  | 1<sup>st</sup> dose WR (Under Pc25) |
|  | 2<sup>nd</sup> dose WR (Under Pc25) |
| **Risk factors** |  |
| Age | Over 75 yo |
| Charlson commorbidity index | Over 8 points |
| Dialysis vintage | Over 24 mo |
| Albumin | Less than 3.5 g/dL |
| iPTH | Less than 150 ng/L |
| C-reactive protein | Over 1 mg/dL |
|  | Sex |
|  | Diabetes |
|  | Vascular access (CVC/AVF) |

AU/mL: arbitrary units per milliliter; NR: Non-responders; WR: Weak-responders; Pc25: Percentile 25; yo: years-old; mo: months; ng/L: nanograms per liter; g/dL: gram per deciliter; mg/dL: milligram per deciliter

For descriptive analysis, continuous variables using means with standard deviations if normal distributed or medians with interquartile range for skewed distribution, whereas categorical variables were presented using as frequencies in with percentages.

For comparative analysis, specific statistical tests for relations between humoral variables and risk factors were performed: 1) Continuous/continuous – Correlation with Spearman for skewed and Pearson for normal distribution; 2) Binominal/Continuous – Differences in median with Mann-Whithney *U* for skewed distribution and means with Student’s t-test if normal distributed; 3) Binominal/Binominal – Fisher’s exact test and Phi coefficient if significant.

The risk factors that showed statistical significance were pooled in for binary regression analysis as predictors to create the best model, associating WR and NR after the 2^nd^ dose of BNT-162b2 with the proposed risk factors.

## RESULTS

Of the forty-six HD patients enrolled, forty-two were eligible for the study: 1) two patients discontinued dialysis before the administration of the second dose; 2) one patient died due to unrelated cause and 3) one patient showed a significant increase in IgM raising the possibility of contact with the virus despite asymptomatic.

### Descriptive analysis

IgG levels achieved after the first dose were low, with a median of 0.99 and 50% rate of NR. After the second dose, median IgG increased to 65.81 and NR rate dropped to 14% (6 patients). The group was mainly composed of elderly patients (mean age 75.1±11.7), with high CCI (7.8±2.8), 59.5 % were female, 45.2 % diabetic, 59.5% used arteriovenous fistula (AVF) as vascular access, while the remaining used a central venous catheter (CVC). Dialysis vintage ranged from 5 to 126 months, with a median of 36 months. Complete descriptive analysis with the remaining demographic, clinical and laboratory variables is summarized in table 2.

**Table 2:** Descriptive analysis of the study variables.

| Continuous | Value | Min/Max | Categorical | N (%) |
| --- | --- | --- | --- | --- |
| Humoral response associated variables |  |  |  |  |
| IgG anti-Spike titers 1<sup>st</sup> dose | 0.99 (3.2) | 0.1/49.2 | 1<sup>st</sup> dose NR | 21 (50) |
|  |  |  | 2<sup>nd</sup> dose NR | 6 (14.3) |
| IgG anti-Spike titers 2<sup>nd</sup> dose | 65.81 (120.56) | 0.1/411.6 | 1<sup>st</sup> dose WR (<0.17 AU/mL) | 10 (23.8) |
|  |  |  | 2<sup>nd</sup> dose WR (<16.29 AU/mL) | 10 (23.8) |
| Risk factors associated variables |  |  |  |  |
| Age, mean (σ) | 75.1 (11.7) | 42/98 | Age, over 75 yo | 24(57.1) |
| Dialysis vintage, mo | 36 (45) | 5/126 | Dialysis vintage, over 24 mo | 24 (57.1) |
| CCI, mean (σ) | 7.8 (2.4) | 3/12 | CCI, over 8 | 23 (54.8) |
| Albumin, mean (σ) | 3.6 (0.5) | 2.2/4.5 | Alb, below 3.5 g/dL | 16 (38.1) |
| CRP | 0.58 (1.08) | 0.12/9.92 | CRP, over 1 mg/dL | 14 (33.3) |
| iPTH | 109.8 (173) | 7/395.5 | iPTH, below 150 ng/L | 25 (59.5) |
|  |  |  | Sex, male | 25 (59.5) |
|  |  |  | Diabetes | 19(45.2) |
|  |  |  | Vascular access, AVF | 25(59.5) |

Continuous variables are presented in median (interquartile range), if not otherwise specified. Min: Minimum value; Max: Maximum value; N (%): Absolute and relative frequency; NR: Non-Responders; WR: Weak Responders; AU/mL: Arbitrary units per milliliter; $\sigma$ : standard deviation; yo: years old; mo: months; CCI: Charlson commorbidity index; Alb: Albumin; CRP: C-Reactive Protein; iPTH: Intact parathormone; AVF: Arteriovenous fistulae; g/dL: gram per deciliter; mg/dL: milligrams per deciliter; ng/L: nanograms per liter.

### Comparative analysis

Table 3 and 4 summarize the relationship between risk factors and humoral variables for the first and second doses, respectively.

**Table 3:**
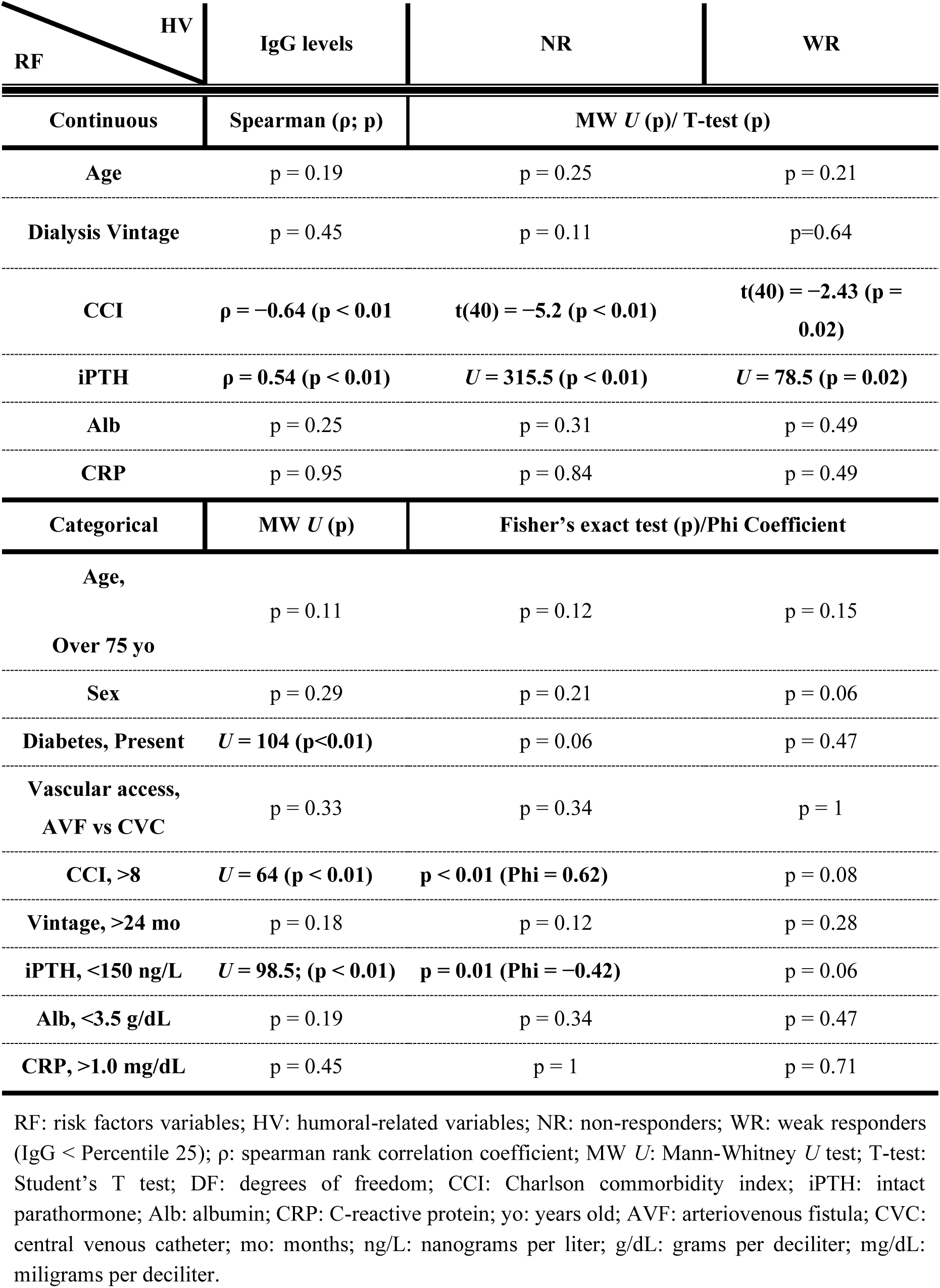
First dose comparative and association analysis between risk-factors and humoral-related variables. Statistically significant values are detailed with additional test-related results.

**Table 4:** Second dose comparative and association analysis between risk-factors and humoral-related variables. Statistically significant values are detailed with additional test-related results.

| RF | HV | IgG levels | NR | WR |
| --- | --- | --- | --- | --- |
| | Continuous | Spearman ($\rho$; p) | MW $U$ ($U$; p)/ T-test (p) | |
| Age | | $\rho = -0.31$ (p = 0.05) | p = 0.33 | t(40) = -2.18 (p = 0.03) |
| Dialysis Vintage |  | p = 0.05 | p = 0.07 | p = 0.07 |
| CCI | | $\rho = -0.66$ (p < 0.01) | p = 0.13 | t(40) = -3.87 (p < 0.01) |
| iPTH | | $\rho = 0.63$ (p < 0.01) | $U = 164.5$ (p = 0.02) | $U = 39.5$ (p < 0.01) |
| Alb | | $\rho = 0.43$ (p < 0.01) | $U = 176$ (p = 0.01) | p = 0.05 |
| CRP |  | p = 0.85 | p = 0.56 | p = 0.61 |
| Categorical | | MW $U$ (p) | Fisher's exact test (p)/Phi Coefficient | |
| Age, Over 75 yo | | $U = 130$ (p = 0.03) | p = 0.214 | p = 0.03 (Phi = 0.37) |
| Sex, female |  | p = 0.09 | p < 0.01 (Phi = -0.495) | p < 0.01 (Phi = 0.45) |
| Diabetes, Present | | $U = 136$ (p = 0.04) | p = 0.67 | p = 0.47 |
| Vascular access, AVF vs CVC | | $U=277$ (p = 0.04) | p = 0.19 | p = 0.01 (Phi = -0.48) |
| CCI, >8 | | $U = 70$ (p < 0.01) | p = 0.67 | p = 0.01 (Phi = 0.40) |
| Vintage, >24 mo |  | p=0.2 | p = 0.07 | p = 0.01 (Phi = -0.42) |
| PTH, <150 ng/L | | $U = 61$ (p < 0.01) | p = 0.07 | p < 0.01 (Phi = 0.45) |
| Alb, <3.5 g/dL | | $U = 120$ (p = 0.02) | p = 0.18 | p = 0.142 |
| CRP, >1.0 mg/dL |  | p=0.42 | p=0.38 | p=1 |

RF: risk factors variables; HV: humoral-related variables; NR: non-responders; WR: weak responders (IgG < Percentile 25); $\rho$ : spearman rank correlation coefficient; MW $U$ : Mann-Whitney $U$ test; T-test: Student's T test; DF: degrees of freedom; CCI: Charlson commorbidity index; iPTH: intact parathormone; Alb: albumin; CRP: C-reactive protein; yo: years old; AVF: arteriovenous fistula; CVC: central venous catheter; mo: months; ng/L: nanograms per liter; g/dL: grams per deciliter; mg/dL: miligrams per deciliter.

#### 1. First dose

IgG levels after the first dose showed a strong negative correlation with CCI (ρ= −0.64, p<0.01) and a positive with iPTH (ρ=0.54, p<0.01). IgG titers were significantly lower for diabetic patients (p<0.01), those with CCI over 8 (p<0.01) and iPTH under 150 ng/L (p<0.01). NR subgroup had a higher CCI and lower iPTH levels (p<0.01 and p<0.01, respectively). Consistently, there was an association with CCI over 8 (Phi=0.62, p<0.01) and iPTH under 150 ng/L (Phi=0.42, p<0.01). WR subgroup, defined as IgG levels under 0.17 AU/mL (Percentile 25), had a higher CCI and lower iPTH levels (p=0.02 and p=0.02, respectively) without any statistically significant association with categorical variables.

#### 2. Second dose

IgG levels after the second dose also showed a strong negative correlation with CCI (ρ= −0.66, p<0.01) and a positive with iPTH (ρ=0.63, p<0.01). A weaker correlation was verified with age (ρ= −0.31, p=0.05) and albumin (ρ=0.43, p<0.01). IgG titers were lower in subgroup analysis for age over 75 years old (p=0.03), diabetic (p=0.04), CVC as vascular access (p=0.04), CCI over 8 (p<0.01), albumin under 3.5 g/dL (p=0.02) and iPTH under 150 ng/L (p<0.01). NR subgroup showed lower albumin (p=0.01) and iPTH levels (p=0.01) and a moderate association with female sex (p<0.01; Phi=0.5). WR subgroup included older patients (p=0.03), with higher CCI (p<0.01) and lower iPTH levels (p<0.01). This subgroup showed multiple associations, namely with vascular access favoring AVF (p=0.01; Phi= −0.48), female sex (p<0.01; Phi=0.45), iPTH under 150 ng/L (p<0.01; Phi= 0.45), dialysis vintage favoring time over 24 months (p=0.01; Phi= −0.42), CCI over 8 (p=0.01; Phi= 0.4) and finally, with the weakest association, age over 75 years (p=0.03; Phi 0.37).

C-Reactive Protein (CRP) showed no significant association or difference in any of the performed statistical analysis.

### Regression analysis

A binominal regression analysis was performed for NR and WR after completing the vaccination schedule using significant risk factors. No model for NR was relevant. A model using CCI (p=0.03), male sex (p=0.02) and CVC as vascular access was statistically significant for predicting WR in this cohort with odds ratio (OR) 1.81, 95% confidence interval (CI) 1.06-3.08, OR 0.05, 95% CI 0.003-0.65 and OR 13.55, 95% CI 1.06-174.18, respectively (p=0.01 for constant).

## DISCUSSION

Despite the well-known association of hemodialysis with a significant immunosuppression state, the overall humoral response to BNT-162b2 vaccine evaluated by IgG anti-Spike titers was significant in our HD study patients, with only 6 patients (14%) failing to achieve a measurable response. It is also important to note that during the follow-up no patient developed clinically significant COVID-19 disease, neither had a PCR-confirmed Sars-CoV-2 infection.

From the beginning, we recognize that our study is limited to the quantification of humoral response, lacking both the assessment of their neutralizing ability and also the associated T cell response, which are required to properly conclude whether the immune response protects against infection or not.^8^ Regarding the method to assess antibody immune response (in arbitrary units), it is important to highlight that its laboratory dependence does not allow for external comparison.

The small sample size, allied with only 6 NR patients after complete vaccination, limited the capacity of this study in properly establish risk factors for NR, which is why the main objective was to study those on risk of low response using the percentile 25 as threshold. Nevertheless, larger studies or pooled analysis are necessary for properly understand which factors influence absence of immunogenicity.

Looking at the demographic results, female sex was associated with lower response. However, a post-hoc analysis of this subgroup showed that women were older and had higher CCI when compared to their male counterparts. Hence, caution should be taken when associating sex and humoral response.

CCI integrates age and diabetes as some of the index’s parameters. Age has been established as a factor for low humoral response in previous studies,^9^ which is also true for ours. Diabetic patients had lower antibody titers, but do not show an association with NR or WR, which was not expected.

When looking at dialysis vintage and vascular access, the analysis of our study population suggested an association between “recently” started HD patients (less than two years) and dialysis by CVC (vs. AVF) with WR. We know that the transition period following HD initiation is unstable and of greatest frailty with decompensation of chronic diseases and physiological changes, which might explain this difference. The use of CVC is associated with more comorbidities and an underling continuous inflammatory response when compared to AVF, which can also compromise the immune system’s response^14^.

Laboratory variables were remarkable in this study. Low iPTH has been previously proposed as a marker of malnutrition and inflammation and established as a poor prognostic factor in HD patients^15-19^. It has also been proposed that a low iPTH is related with immunologic dysfunction, with a higher risk of infection in ESRD patients^19^. In this study, lower iPTH levels were consistently associated with an overall low humoral response. Every patient in the WR and NR subgroups had iPTH under 150 ng/L. In a similar way, albumin, both as a marker of malnutrition or negative inflammation marker,^20-22^ shows a negative association with humoral variables, particularly after completing the vaccination schedule. CRP did not show any statistical significance, which was unexpected, since inflammation is known to affect immunogenicity.

Despite the undeniable requirement for further investigation, this study may be the starting point for the definition of a risk stratification panel in which multiple factors are key elements for the identification of patients at risk of an unsatisfactory response to vaccination. Their identification may be crucial, providing institutions the opportunity to adapt contingency protocols.

## CONCLUSION

Quantifiable humoral response to BNT-162b2 is significative among HD patients.

Our study also suggests a patient-centered evaluation for risk of low immunogenicity using various surrogate markers, namely CCI, PTH, albumin and type of vascular access. Age alone, as established by other studies, can be considered, but might be insufficient when compared to a more complete comorbidity assessment. A patient-centered risk stratification for low immunogenicity using a risk factor panel, rather than age alone, is proposed as a better surrogate marker for immunogenicity.

Proper identification of these high-risk patients for low to absent response may play a key role in policy creation and adjustment while providing insight for proper contingency planification. Closer monitoring, adequate isolation, vigilance of symptoms and identification of those that may benefit from booster doses are some of the potential advantages of using this holistic approach.

A larger study enrolling more patients and possible introducing the evaluation of cellular immunization is warranted to firmly establish high-risk patients in order to create the best surrogate model for low immunogenicity in this population.

Widespread vaccination of the HD population remains the single most important form of prevention of COVID-19 and should be expeditiously instituted worldwide.

## Data Availability

The data present in this work is only available in clinical processes and is not of public domain or access. All information is under patient consent access.

## Author contributions

Concept and design: Rui Duarte, Francisco Ferrer, Ivan Luz

Acquisition and interpretation of data: Rui Duarte, Cátia Figueiredo

Drafting of the manuscript: Rui Duarte

Intellectual revision of content: Ivan Luz, Francisco Ferrer

Statistical analysis: Hernâni Gonçalves

Language and final revision: Francisco Ferrer, Karina Lopes, Ivan Luz

Technical and administrative support: Flora Sofia

Supervision and approval of the article: Karina Lopes

---

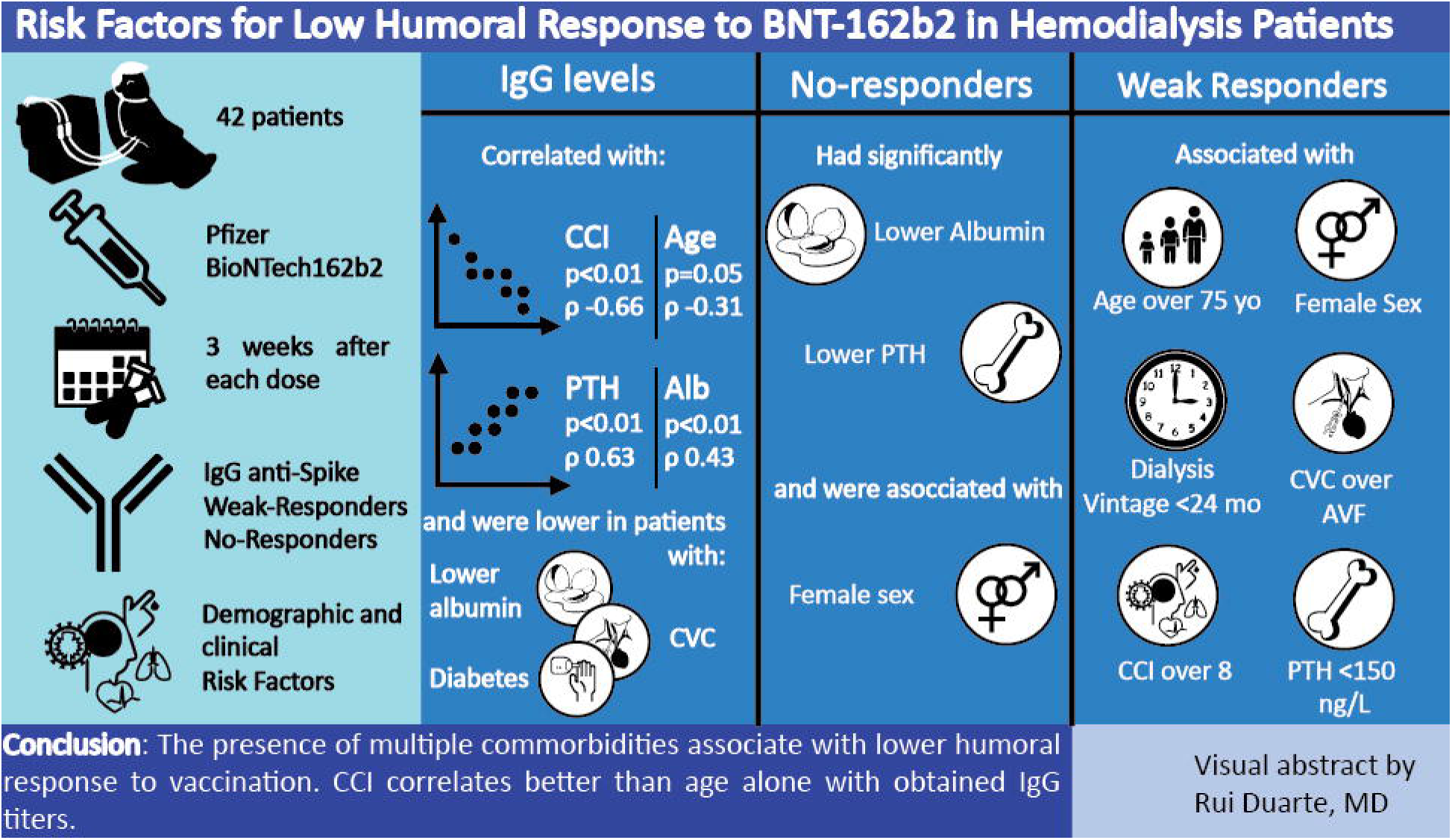

## REFERENCES

1. Polack FP, Thomas SJ, Kitchin N, et al. Safety and Efficacy of the BNT162b2 mRNA Covid-19 Vaccine. N Engl J Med. 2020;383(27):2603–2615

2. Anderson EJ, Rouphael NG, Widge AT, et al. Safety and Immunogenicity of SARS-CoV-2 mRNA-1273 Vaccine in Older Adults. N Engl J Med. 2020;383(25):2427–2438

3. Ramasamy MN, Minassian AM, Ewer KJ, et al. Safety and immunogenicity of ChAdOx1 nCoV-19 vaccine administered in a prime-boost regimen in young and old adults (COV002): a single-blind, randomised, controlled, phase 2/3 trial. Lancet (London, England). 2021;396(10267):1979–1993.

4. Stephenson KE, Le Gars M, Sadoff J, et al. Immunogenicity of the Ad26.COV2.S Vaccine for COVID-19. JAMA. 2021;325(15):1535–1544

5. Wu C, Chen X, Cai Y, et al. Risk Factors Associated with Acute Respiratory Distress Syndrome and Death in Patients with Coronavirus Disease 2019 Pneumonia in Wuhan, China. JAMA Intern Med. 2020;180(7):934–943

6. Flythe JE, Assimon MM, Tugman MJ, et al. Characteristics and Outcomes of Individuals with Pre-existing Kidney Disease and COVID-19 Admitted to Intensive Care Units in the United States. Am J kidney Dis Off J Natl Kidney Found. 2021;77(2):190–203

7. Valeri AM, Robbins-Juarez SY, Stevens JS, et al. Presentation and Outcomes of Patients with ESKD and COVID-19. J Am Soc Nephrol. 2020;31(7):1409–1415.

8. Broseta JJ, Rodríguez-Espinosa D, Rodríguez N, Mosquera MDM, Marcos MÁ, Egri N, Pascal M, Soruco E, Bedini JL, Bayés B, Maduell F. Humoral and Cellular Responses to mRNA-1273 and BNT162b2 SARS-CoV-2 Vaccines Administered to Hemodialysis Patients. Am J Kidney Dis. 2021 Jun 24:S0272-6386(21)00689

9. Grupper A, Sharon N, Finn T, et al. Humoral Response to the Pfizer BNT162b2 Vaccine in Patients Undergoing Maintenance Hemodialysis. Clin J Am Soc Nephrol. Published online 2021

10. Attias P, Sakhi H, Rieu P, et al. Antibody Response to the BNT162b2 Vaccine in Maintenance Hemodialysis Patients. Kidney Int. 2021;99(6):1490–1492.

11. Ikizler TA, Coates PT, Rovin BH, Ronco P. Immune Response to SARS-CoV-2 Infection and Vaccination in Patients Receiving Kidney Replacement Therapy. Kidney Int. 2021;99(6):1275–1279

12. Stevens CE, Alter HJ, Taylor PE, Zang EA, Harley EJ, Szmuness W. Hepatitis B Vaccine in Patients Receiving Hemodialysis. N Engl J Med. 1984;311(8):496–501.

13. Buti M, Viladomiu L, Jardi R, et al. Long-Term Immunogenicity and Efficacy of Hepatitis B Vaccine in Hemodialysis Patients. Am J Nephrol. 1992;12(3):144–14

14. K. Krueger, M. Ison, C. Ghossein. Practical Guide to Vaccination in All Stages of CKD, Including Patients Treated by Dialysis or Kidney transplantation. Am J Kidney Dis, 75 (2020), pp. 4176–4425

15. Hung AM, Ikizler TA. Hemodialysis central venous catheters as a source of inflammation and its implications. Semin Dial. 2008 Sep-Oct;21(5):401–4.

16. Avram MM, Mittman N, Myint MM, Fein P. Importance of low serum intact parathyroid hormone as a predictor of mortality in hemodialysis and peritoneal dialysis patients: 14 years of prospective observation. Am J Kidney Dis. 2001 Dec;38(6):1351–7.

17. Yu Y, Diao Z, Wang Y, et al. Hemodialysis patients with low serum parathyroid hormone levels have a poorer prognosis than those with secondary hyperparathyroidism. Ther Adv Endocrinol Metab. 2020 Sep 21, vol. 11:2042018820958322.

18. D. Angelini, A. Carlini, R. Giusti et al., “Parathyroid hormone and T-cellular immunity in uremic patients in replacement dialytic therapy,” Artificial Organs, vol. 17, no. 2, pp. 73–75, 1993.

19. C. Tzanno-Martins, E. Futata, V. Jorgetti, and A. J. S. Duarte, “Restoration of impaired T-cell proliferation after parathyroidectomy in hemodialysis patients,” Nephron, vol. 84, no. 3, pp. 224–227, 2000.

20. Fukagawa, Masafumia; Akizawa, Tadaob; Kurokawa, Kiyoshic Is aplastic osteodystrophy a disease of malnutrition?, Current Opinion in Nephrology and Hypertension: July 2000 - Volume 9 - Issue 4 - p 363–367

21. Cohen G, Hörl WH. Immune dysfunction in uremia—an update. Toxins (Basel). 2012;4(11):962–990.

22. Gama-Axelsson T, Heimbürger O, Stenvinkel P, et al. Serum albumin as predictor of nutritional status in patients with ESRD [published correction appears in Clin J Am Soc Nephrol. 2012 Nov;7(11):1915]. Clin J Am Soc Nephrol. 2012;7(9):1446–1453.

